# BENEFITS OF PALLIATIVE CARE IN ADULTS WITH A DIAGNOSIS OF HEART FAILURE: AN EXPLORATORY LITERATURE REVIEW

**DOI:** 10.1101/2024.04.01.24305119

**Authors:** Aura Cristina Herrera Obando, Jhaina Isabella Larrañaga Guerrero, William Antonio Mejia Montehermoso, Simon Andres Giraldo Oliveros

## Abstract

**Introduction:** Heart Failure is a clinical syndrome characterized by a series of symptoms such as dyspnea, orthopnea and edema in the lower limbs. This pathology continues to have a high prevalence despite advances in pharmacotherapy and device therapy and given that it is a pathology that significantly impairs the quality of life of patients, the implementation of care is of vital importance. However, these are underused due to lack of knowledge on the part of health personnel and also due to poor implementation in the different health providers.

**Objective:** An exploratory review of the literature was carried out regarding the benefits of palliative care in patients with advanced heart failure, in order to synthesize the available and updated evidence.

**Methodology:** Searched for articles published from 2017 to 2022 related to palliative care in patients with heart failure and using the PRISMA 2020 methodology for this study. This inquiry of articles was carried out in the following databases: UpToDate, PubMed, MESH, PMC (US National Library of Medicine National Institutes of health).

**Results:** A total of 5 articles were obtained, from which they concluded that palliative care has a positive impact on the quality of life of patients with heart failure, there was a lower rate of hospital readmissions, improvements in physical, psychological and existential.

## INTRODUTION

Heart failure (HF) is a clinical syndrome characterized by a group of symptoms (dyspnea, orthopnea, lower limb edema) and signs (elevated jugular venous pressure, pulmonary congestion) that are often caused by structural and/or functional cardiac abnormalities resulting in decreased heart rate, cardiac output and/or elevated intracardiac pressures (1).

Despite multiple advances in pharmacotherapy and device therapies, heart failure continues to progress, becoming a highly symptomatic and fatal pathology that places great demands on patients, caregivers, health care professionals such as physicians, nurses and others, and the health care system in general (2).

Prevalence and incidence continue to increase; the most recent estimates for 2015 indicate a worldwide prevalence of 23 million patients with heart failure (3). According to projections by the Economic Commission for Latin America and the Caribbean (ECLAC), by 2030 in Colombia the prevalence of patients with heart failure will have increased to 18.6%, reaching 27.4% of patients with heart failure by 2050 (4).

Patients with heart failure demand palliative care with a multidisciplinary approach focused on the control of their physical and psychosocial symptoms in order to better improve the quality of life of both patients and their families, through the prevention and relief of suffering, thanks to early identification, evaluation and adequate treatment of pain and other physical, psychosocial and spiritual problems (5). Although there are studies that support the implementation of palliative care, it is underutilized since it is implemented only in advanced stages of pathology or in the hospital setting and is rarely used proactively in outpatients and in early stages of heart failure (6).

This exploratory literature review aims to synthesize the evidence of the benefits of palliative care in order to determine how it improves the quality of life of patients and their families, so that it can be implemented as an early and effective tool within conventional treatment.

## METHODOLOGY

A systematic review of the literature is performed, through an active search of review articles and clinical trials published between 2017 and 2022 in databases such as PubMed, Lilacs, Google Scholar, ScienceDirect and Scopus. Articles in Spanish and English were included. The following MeSH and DeCS terms were used for the search: Heart Failure, Palliative care and Life Quality. The search yielded 1068 articles, of which 1063 were excluded for duplicity or for not meeting the inclusion criteria or objectives of this review. This article is based on 5 articles that met the search criteria and were registered in indexed journals.

## RESULTS

For the development of the exploratory review, the following databases and search engines were used (PubMed, Science direct, Google academic, Lilac and Scopus), in which the main strategy used were keywords that were chosen and reviewed in the MeSH and DeCS platforms (“Heart failure” AND “Palliative care” AND “Life Quality”) and a selection of time period established from the year 2017 to 2022, thus providing us with a total of 1068 studies, of which 442 were excluded due to the fact that repeated articles were found in the platforms consulted, thus leaving us with 626. A pre-selection of each search source was made, where studies that spoke about the topic to be explored were chosen in the previous summary of each one, resulting in 36 articles to be read; as a third step, a second, more exhaustive pre-selection was made in which the introduction of each article, type of study and conclusions were taken into account, in which 16 of them were selected to begin with the complete selection (Figure 1).

**Figure 1.**
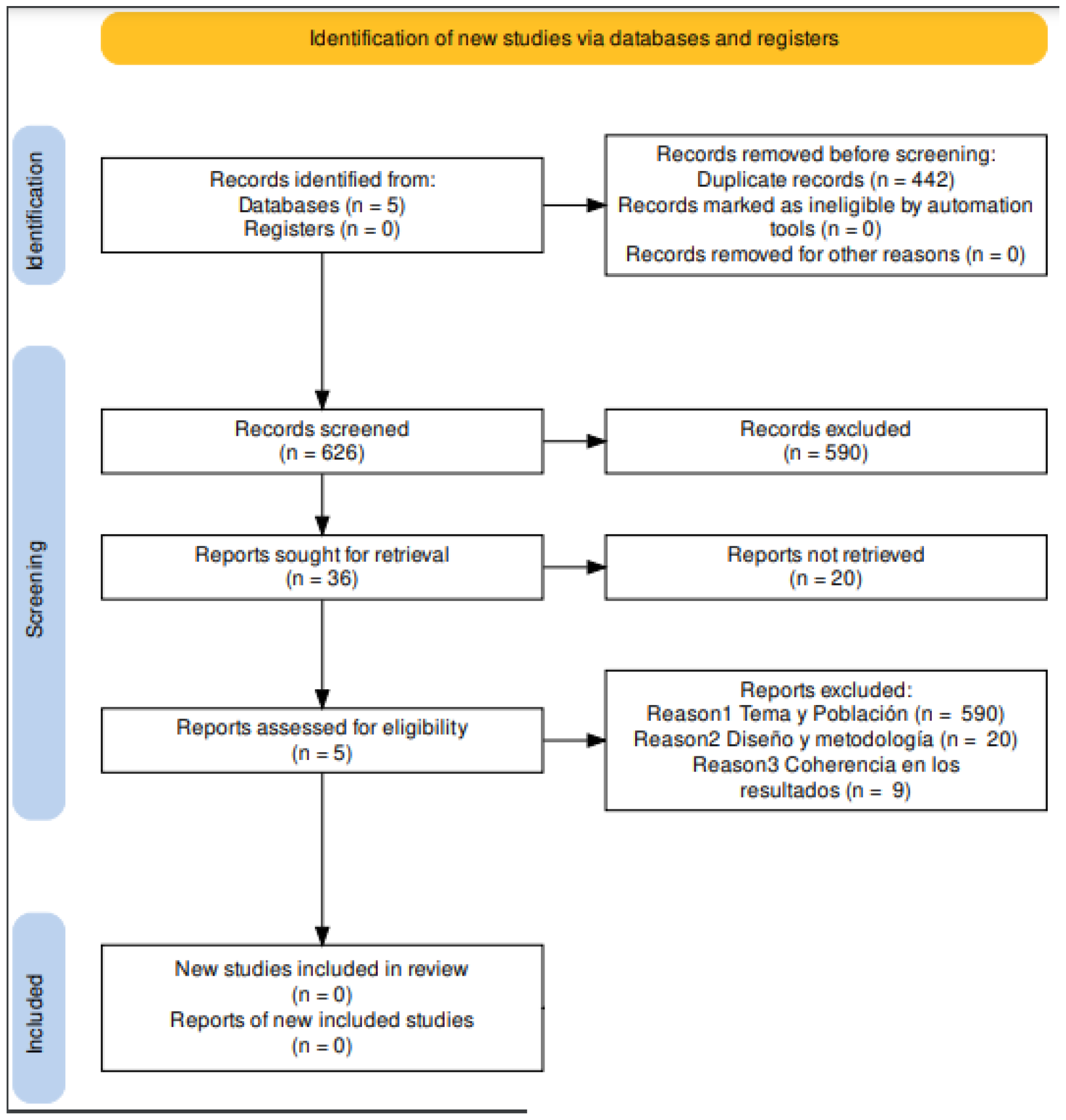
PRISMA flowchart for the selection of the body of evidence

The body of evidence selected on the benefits of palliative care in patients diagnosed with advanced heart failure was written in only two countries, with a higher frequency of scientific production in the United States of America (80% of the total articles) and the rest in the People“s Republic of China (20% of the total articles); among the evidence collected, it was not found that any author participated in more than one study; Within the universities in which the studies were developed, 100% of them correspond to medical programs which are characterized by having high quality research programs, achieving that they are highlighted by being within the list of the best 10 universities nationally and worldwide, and thus obtaining accreditations such as the Liaison Committee on Medical Education (LCME) and the Center for Medicare and Medicaid Services, which provide clinics and hospitals of medium and high complexity. Within the temporal range of publication was between the years 2017 to 2020, where curiously in the years 2017 and 2018 reported a greater number of publications that were disseminated in the best and most prominent journals related to internal medicine and cardiology, finding that in scimago was performed an exhaustive search of each journal from where the publications were made evidencing:

The first is the Journal of the American College of Cardiology, a leader in the field of cardiovascular disease, which focuses on drug therapy, new diagnostic techniques, laboratory discoveries and large multicenter studies of new treatments, has an H index of 451 and has been published since 1983; Journal of Pain and Symptom Management is a journal that publishes the latest clinical research and best practices related to reducing the burden of illness in critically ill patients, has been a strong advocate of quantitative and qualitative research that supports the development of the palliative care discipline, has an H-index of 147 and has been published since 1986; JAMA Cardiology is a journal focused on all aspects of cardiovascular medicine, prevention, diagnostic testing, interventions, and drug therapies, has an H-index of 80 and has been published since 2016; JAMA internal medicine provides research innovative and clinically relevant for general internists and internal medicine subspecialties. Articles published in this journal have resulted in FDA actions and congressional hearings on patient safety issues with the goal of taking better steps to protect the public, it has an Hindex of 358 and has been published since 2013; and finally the Journal of the American Heart Association provides a worldwide forum for basic and clinical research articles and timely reviews on cardiovascular disease and stroke, has an H-index of 100 and has been published since 2012 (Table 4).

**TABLE 4.** Characteristics of the publication of articles

| N° | AUTHOR | COUNTRY | LANGUAGE | UNIVERSITY | HOSPITAL | YEAR OF PUBLICATION | JOURNAL OF PUBLICATION | INDEX H OF THE JOURNAL | QUARTILE OF THE JOURNAL |
| --- | --- | --- | --- | --- | --- | --- | --- | --- | --- |
| 1 | Joseph G. Rogers MD | United States (Durham, North Carolina) | English | Duke University School of Medicine | Duke Clinical, research institute | 2017 | Journal of the American College of Cardiology | 451 | Q1 |
| 2 | Alina Yee Man | China (Hong Kong) | English | The Hong Kong Polytechnic University | Grantham Hospital, Heaven of hope Hospital, United Christian Hospital. | 2017 | Journal of Pain and Symptom Management | 147 | Q1 |
| 3 | Arden E. O'Donnell | Boston, Massachusetts | English | Boston University, Boston, Massachusetts | Brigham and Women's Hospital | 2018 | JAMA cardiology | 80 | Q1 |
| 4 | Bekelman DB | Denver, Colorado | English | Universidad de Colorado, Campus Médico Anschutz, Aurora | NA | 2018 | JAMA internal medicine | 358 | Q1 |
| 5 | Michelle S. Diop | United States | English | Medical School of Brown University | Center of Innovation in Long-Term Services and Supports, Providence VA Medical Center | 2020 | Journal of the American Heart Association | 100 | Q1 |

**TABLE 5.** Characteristics of the study design

| N° | AUTHOR | TYPE OF STUDY | N° OF PARTICIPANTS | AVERAGE AGE | % MEN | % WOMAN |
| --- | --- | --- | --- | --- | --- | --- |
| 1 | Joseph G. Rogers MD | Prospective, single-site randomized controlled trial | 150 | 70,9 | 53% | 47% |
| 2 | Alina Yee Man | Randomized controlled trial | 84 | 78,4 | 51,1% | 48.9% |
| 3 | Arden E. O'Donnell | Randomized Clinical Trial | 50 | 72 | 58% | 42% |
| 4 | Bekelman DB | Multicenter randomized clinical trial | 317 | 65,5 | 78,70% | 21,30% |
| 5 | Michelle S. Diop | Retrospective Paired Propensity Analysis | 2862 | 75,7 | 98,6 % | 1,40% |

Within the articles selected for the body of evidence the number of participants ranged between 50 and 317, most of them corresponded to clinical trials, by type of disease the average age is above 65 years, the study population mostly corresponds to men, with an average age in general of 70 years old, it is noteworthy that most of the studies are clinical trials, Each study was carried out in a different country, and the population was taken from different places, i.e. hospitals, clinics, geriatric homes or even some patients were at home where their evolution with palliative care was observed.

**Table 6.**
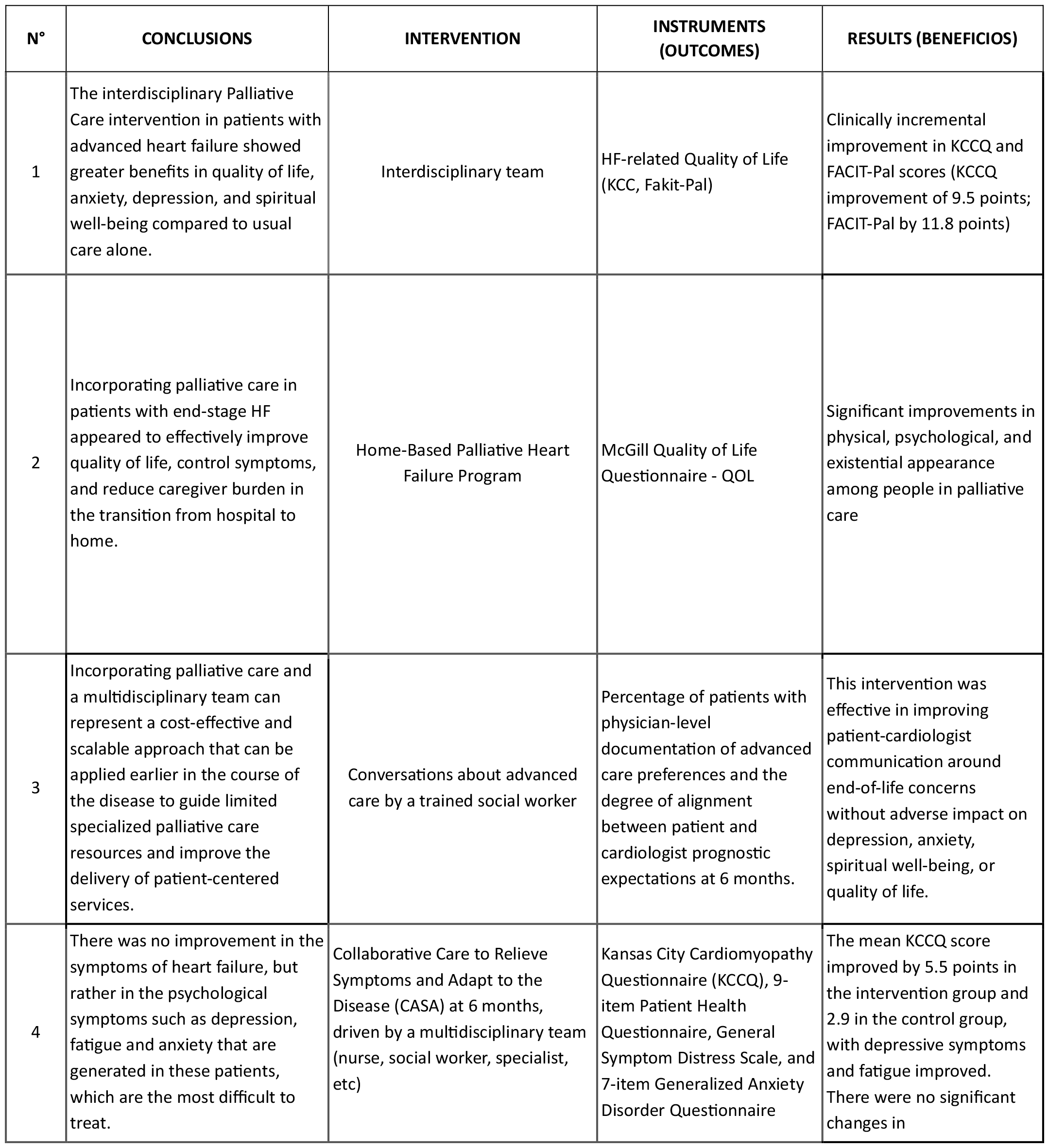
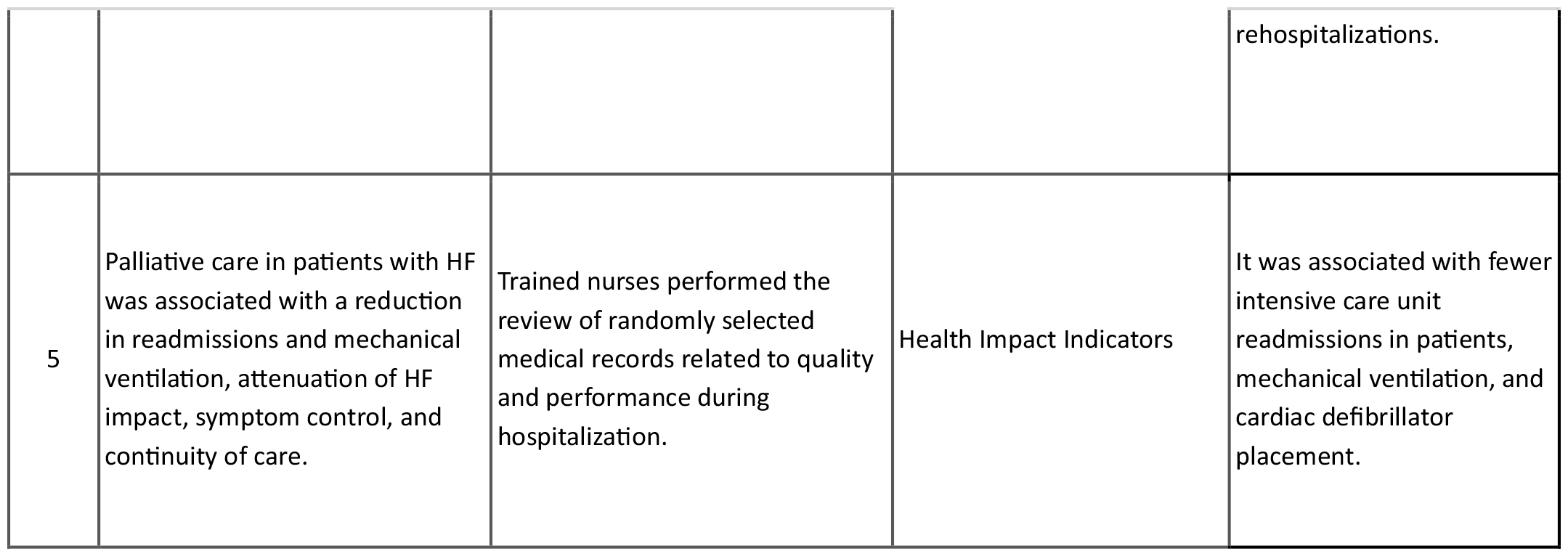
Research results

| N° | CONCLUSIONS | INTERVENTION | INSTRUMENTS (OUTCOMES) | RESULTS (BENEFICIOS) |
| --- | --- | --- | --- | --- |
| 1 | The interdisciplinary Palliative Care intervention in patients with advanced heart failure showed greater benefits in quality of life, anxiety, depression, and spiritual well-being compared to usual care alone. | Interdisciplinary team | HF-related Quality of Life (KCC, Fakit-Pal) | Clinically incremental improvement in KCCQ and FACIT-Pal scores (KCCQ improvement of 9.5 points; FACIT-Pal by 11.8 points) |
| 2 | Incorporating palliative care in patients with end-stage HF appeared to effectively improve quality of life, control symptoms, and reduce caregiver burden in the transition from hospital to home. | Home-Based Palliative Heart Failure Program | McGill Quality of Life Questionnaire - QOL | Significant improvements in physical, psychological, and existential appearance among people in palliative care |
| 3 | Incorporating palliative care and a multidisciplinary team can represent a cost-effective and scalable approach that can be applied earlier in the course of the disease to guide limited specialized palliative care resources and improve the delivery of patient-centered services. | Conversations about advanced care by a trained social worker | Percentage of patients with physician-level documentation of advanced care preferences and the degree of alignment between patient and cardiologist prognostic expectations at 6 months. | This intervention was effective in improving patient-cardiologist communication around end-of-life concerns without adverse impact on depression, anxiety, spiritual well-being, or quality of life. |
| 4 | There was no improvement in the symptoms of heart failure, but rather in the psychological symptoms such as depression, fatigue and anxiety that are generated in these patients, which are the most difficult to treat. | Collaborative Care to Relieve Symptoms and Adapt to the Disease (CASA) at 6 months, driven by a multidisciplinary team (nurse, social worker, specialist, etc) | Kansas City Cardiomyopathy Questionnaire (KCCQ), 9-item Patient Health Questionnaire, General Symptom Distress Scale, and 7-item Generalized Anxiety Disorder Questionnaire | The mean KCCQ score improved by 5.5 points in the intervention group and 2.9 in the control group, with depressive symptoms and fatigue improved. There were no significant changes in rehospitalizations. |
| 5 | Palliative care in patients with HF was associated with a reduction in readmissions and mechanical ventilation, attenuation of HF impact, symptom control, and continuity of care. | Trained nurses performed the review of randomly selected medical records related to quality and performance during hospitalization. | Health Impact Indicators | It was associated with fewer intensive care unit readmissions in patients, mechanical ventilation, and cardiac defibrillator placement. |

Of the four clinical trials it is noteworthy that the first article in the table authored by Josep G and Rodgers MD focused on the investigation of interdisciplinary palliative care management compared to usual heart failure care, this study was based primarily on patients with advanced heart failure, with high mortality risk and a life expectancy of six months, where there were two fundamental endpoints to measure the quality of HF (Kansas City Cardiomyopathy Questionnaire [KCCQ]) and health-related quality of life (Functional Assessment of Chronic Illness Therapy-Palliative Care [FACIT-Pal] scale); while the second article authored by Alina Yee Man examined the effect of a home-based palliative heart failure (HPHF) program on quality of life (QOL), symptom burden, functional status, patient satisfaction and caregiver burden among patients with ESHF and performed using the McGill-Hong Kong Quality of Life Questionnaire; the third trial, authored by Arden E. O“Donnell, randomly selected 50 patients who were hospitalized, and the intervention evaluated was the consultation with a social worker with five years of experience in palliative care and the control was to follow the standard or usual care, in order to compare the benefits of this consultation. What the social worker sought was to talk to the patient in order to analyze the prognosis of their disease and resolve all doubts in order to guide patients in understanding their prognosis, preferences at the end of life, evaluating depression, anxiety, spiritual well-being or quality of life; however, this was not significant because it was not evaluated whether it improved their prognosis or worsened it, that is to say, this study is inconclusive in quantitative terms.

It should be noted that although the three previous trials were evaluated by different scales, their objective was to measure physical function, symptoms, social function and quality of life in both of them, although a difference was found in the population size, as in the Joseph G. Rogers MD trial which recruited 150 patients and the Alina Yee Man trial which worked with 84 patients, it is considered that they are perfectly comparable given that there is not much difference. Rogers MD who recruited 150 patients and that of Alina Yee Man who worked with 84 patients, it is considered that they are perfectly comparable given that the difference is not much and in both excellent results were obtained in palliative care, and above all highlighting that they were performed in the outpatient setting, i.e. care was provided at home, although there were approximately 4 patients who had to be rehospitalized due to complications, However, this did not greatly affect the studies since it was possible to prove that even though advanced heart failure imposes physical, psychosocial and spiritual burdens, these significantly improve both the patients and their families, as long as they are managed comprehensively and from home, compared to the third clinical trial authored by David B. Bekelman, a multicenter trial. Bekelman, multicenter type, which has the advantage of a faster recruitment, however, they used the same scale of the first article [KCCQ], where they intervened in collaborative psychosocial and symptom care to improve the specific health status of heart failure, depression and symptom burden (pain, shortness of breath, fatigue or depression) in patients with heart failure, One of the reasons may be that the study was carried out in an in-hospital setting where the patients will feel a little uncomfortable due to their condition, i.e. they will be depressed and stress is a triggering factor for the patient to worsen their life condition and make it more difficult for them to adhere to the treatment, especially in this case, which is carried out by a multidisciplinary team and requires constant accompaniment.

In conclusion, and with the results obtained by applying the scales mentioned above, palliative care does improve the quality of life of the patients, from the physical, psychosocial, spiritual, symptoms, depression, etc. aspects. This care is applied by the respective multidisciplinary team.

## DISCUSSION

The average age of the patients in the study was mostly above 70 years, this is explained by the fact that the incidence of the disease increases with each decade of life, which is confirmed by several studies that have shown that heart failure is the main cause of hospitalization in people over 65 years of age, given that the social and health impact is aggravated, associated with high rates of disability and mortality, as a result of physiological deterioration due to cardiovascular aging in which progressive structural damage of the myocardial fibers is evident (25).

According to the collection of data from the articles on which this research was based, a high prevalence of the disease was found in male patients with a large difference compared to the female sex (65%-78%), similar to that of the JACC study and the Spanish journal of cardiology in which they reported that 68% of patients suffering from this disease are men. However, it has been shown that although the prevalence is higher in men, they are less likely to suffer from this disease symptomatically, unlike women who stand out for suffering early symptoms during their pathology (26).

To our knowledge, this exploratory literature review is the first in Colombia, which is based on high impact Q1 journals with high H index (80-451) where the results were collected, which makes it a reliable study for readers and for future studies. Although the sources of information and the articles selected were scarce, they are based on databases that are updated periodically and have been recognized as leaders in the field of clinical reports on cardiovascular diseases.

Previous studies on palliative care interventions in patients with heart failure are scarce, especially in our setting because most have focused on providing conventional treatments without extending multidisciplinary in-hospital and out-of-hospital care in order to provide the patient with more humanized management and ongoing support for the patient and family. Therefore, we want to demonstrate through this review the benefits of implementing this care since it has been shown that there is a great impact on the improvement of the quality of life in these patients, in order to provide an important guide for the clinical practice of health personnel (4,27). Our results add to a current body of knowledge on the effectiveness of a new therapeutic model of PC that requires fewer resources for support and implementation in out-of-hospital HF patients, as well as expand an evolving medical literature that emphasizes the implementation of different treatment strategies and approaches to patients with cardiovascular disease from the time of diagnosis to the end of pathology, which will serve as training and new knowledge acquisition for both trainees and specialized health care personnel.

Our findings have supported the benefits of PC as an intervention capable of improving quality of life in HF patients physically, psychosocially, and spiritually, demonstrating increased patient satisfaction due to reduced use of health services and a sense of reduced caregiver burden, as well as improved communication between the patient and cardiologist about end-of-life concerns without an adverse impact on the patient“s depression, anxiety, and spiritual well-being. Palliative care is an additional utility that cardiologists can use in the comprehensive management of HF patients instead of providing conventional, non-humanized, multidisciplinary treatment that does not take into account different variables or domains of each human being. The comparison observed with reduced re-hospitalization within 6 months among patients who received palliative care may provide additional support that a palliative approach can be used to help guide the goals of care practices for patients living with HF (28).

Among the multiple benefits of the implementation of PCs it has been found that they have been effective in improving communication between the patient and the cardiologist around end-of-life concerns, this demonstrated that there had been reported failures in communication between the treating physician and the patient, establishing that patients with heart failure often tend to underestimate the severity of their disease process and overestimate their own life prognosis, thus having survival expectations that discrepancy considerably from the usual anticipated evolution of their disease (29).

As mentioned above, palliative care is not well established worldwide, and with the literature review in many countries there is an attempt to integrate these programs involving primary care setting and collaboration with general practitioners, where there must be a coordination of care, However, each patient has a different evolution and not all are exempt from rehospitalization, and although the cardiologist plays a very important role in the palliative care of patients with heart failure, it is possible to work together with the specialists of the hospital and the general practitioners in the palliative care of patients with heart failure, The cardiologist plays a very important role in palliative care in patients with heart failure, and although the cardiologist plays a very important role in palliative care in patients with heart failure, you can work together with oncology specialists who have extensive experience in terminally ill patients and undoubtedly palliative care specialists, taking into account that in a quality care not only acts the treating physician, but should be performed by a multidisciplinary team, an example of this I know was found in the literature in the UK where the British Heart Foundation has recently funded where HF nurses who will receive specific training in palliative care and advanced communication skills. This should improve support for both patients and their families and facilitate coordination with other health professionals. A second example of this model is found in Switzerland, where community-based mobile palliative care teams provide home-based patient symptom management and support families in their caregiving role (30).

Some recommendations should be mentioned in the care that is provided with quality where patients and their families are involved, in this long process it is oriented as is the treatment, the wills of life, the objectives of care should be evaluated continuously during the progression of the disease, the conversation with patients and families should focus on what is sought is to improve the quality of life (31-33). It is noteworthy that in the studies reviewed there is uncertainty in palliative care, because not all patients assimilate it satisfactorily so there should be more alternatives to provide patients, and an important point that I observed was depression that although in this care is sought to avoid, many fall into depression due to multiple circumstances, This increases the mortality rate considerably, so in many occasions health professionals face a great challenge, however when depression treatments are provided, there is no evidence that reduces morbidity and mortality, at this point it is extremely important the spiritual satisfaction that can modify the depression state positively, which in its absence improves the quality of life (33).

Palliative care focuses on improving the patient“s quality of life by controlling pain and other distressing symptoms of a serious illness that are disabling for them and have a negative impact on the social, spiritual, physical and mental aspects of their life and that of their caregiver. Palliative care can be provided in conjunction with other medical treatments, with the aim of minimizing symptoms that cause suffering and increasing quality of life (33-35).

### Limitations

One of the main limitations of the present study was the scope of the design, which was only exploratory and did not amount to a systematic review of the benefits of palliative care in patients with heart failure; however, most of the methodological aspects followed the Cochrane Collaboration“s manual for systematic reviews.

It is considered that a critical reading should have been done as CASPe of the selected body of evidence; however, this aspect was attempted to be made up for with the complete reading of the article taking into account the STROBE checklist of the articles.

Another limitation corresponds to the fact that the differential of progression or chronicity of heart failure was not analyzed, which is a relevant factor when evaluating the benefits of palliative care; however, this was approached from the qualitative synthesis, although the studies did not allow this aspect to be clearly established.

## CONCLUSIONS

- Although the literature search yielded many results, when evaluating the quantified impact of palliative care we found a significant reduction, finding many investigations with a spiritual and holistic approach, which, although they contribute to the knowledge of the benefits of palliative care, are not conclusive.
- The sheer volume of scales to measure quality of life and other health-related quality of life benefits of palliative care interventions in heart failure patients does not allow for a standard in comparability, finding that the most commonly used scale is the Kansas City Cardiomyopathy Questionnaire to measure health-related quality of life.
- This exploratory literature review showed that the implementation of palliative care as an adjunct to conventional treatment has a great impact on improving the quality of life of patients diagnosed with HF, thus improving the bothersome symptoms of the disease, mental health and an improvement in terms of the social environment in the life of each patient, in addition to reducing the emotional and physical burden of the caregiver in charge.

## Data Availability

All data produced in the present work are contained in the manuscript

